# Prevalence of traumatic psychological stress reactions in children and parents following paediatric surgery: a systematic review and meta-analysis

**DOI:** 10.1101/2021.04.21.21255853

**Authors:** David Turgoose, Stephanie Kerr, Paolo De Coppi, Simon Blackburn, Simon Wilkinson, Natasha Rooney, Richard Martin, Suzanne Gray, Lee D Hudson

## Abstract

**Background:** Children undergoing surgery, and their parents, are at risk of developing post-traumatic stress reactions. We systematically reviewed the literature to understand the prevalence of this issue, as well as potential risk factors.

**Methods:** We conducted a systematic review and meta-analysis, using PubMed, PsycInfo, Web of Science and Google Scholar, with searches conducted in February 2021. Papers were included if they measured post-traumatic stress in children and/or parents following paediatric surgery, and were excluded if they did not use a validated measure of post-traumatic stress. Data was extracted from published reports.

**Findings:** Our search yielded a total of 1,672 papers, of which 16 of met our inclusion criteria. In meta-analysis, pooled studies of children estimated an overall prevalence of 16% meeting criteria for post-traumatic stress disorder post-surgery (N= 187, 95% CI : 5-31%, I^2^ = 80%). After pooling studies of parents, overall prevalence was estimated at 23% (N= 1444, 95% CI 16-31%, I^2^ = 91%). Risk factors reported within studies included length of stay, level of social support and parental mental health.

**Interpretation:** There is consistent evidence of traumatic stress following surgery in childhood which warrants further investigation. Those delivering surgical care to children would benefit from a raised awareness of the potential for post-traumatic stress in their patients and their families, including offering screening and support.

**Funding:** We did not seek or receive any funding for this study.

## Introduction

Traumatic psychological stress reactions can develop in individuals who experience frightening or life-threatening events.^1^ Post-traumatic stress reactions incorporate a range of psychological symptoms. Diagnostic criteria exist to capture these experiences, namely in the form of Post-Traumatic Stress Disorder (PTSD) and Acute Stress Disorder (ASD). Symptoms of PTSD and ASD include re-experiencing, avoidance, hyperarousal, and negative alterations in cognitions and mood.^2-4^

Research suggests that lifetime exposure to at least one traumatic event is quite common at around 90%.^5^ Lifetime prevalence of PTSD in adults has been estimated at 6.8%^6^ to 9.8%.^5^ In children, studies indicate that 14 - 43% of children and adolescents are exposed to at least one traumatic event in their lifetime, and that up to 15% go on to develop PTSD, with girls being at higher risk than boys.^7^ It is clear therefore that psychological trauma is a significant issue in both child and adult populations.

PTSD has been linked to a range of psychological and interpersonal difficulties, including comorbid mental health difficulties such as depression, substance misuse, suicidal ideation,^8^ eating disorders,^9^ anxiety disorders,^10^ and poor family relationships.^11^ Given that traumatic stress reactions can have such an impact, it is important that services can effectively identify and support families in cases where they may be exposed to trauma. Research is limited, however, and there is a lack of understanding of the risk that surgery poses in terms of the development of post-traumatic stress.

Emerging evidence suggests that children undergoing surgery are at risk of developing traumatic stress reactions post-surgery,^12-15^ as are parents of children undergoing surgery.^16,17^ To date, there is only sporadic evidence of, and no consensus on, the rates of traumatic stress development following surgery, or if any risk factors, such as parental stress,^18^ might increase the chances of such difficulties occurring.

An understanding of post-traumatic stress following surgery in childhood is important, to allow for opportunities to mitigate risks or provide appropriate psychological support. Although there is emerging evidence, the literature on this topic has not yet been systematically synthesised. We therefore conducted a systematic review and meta-analysis using PRISMA guidance of the literature on 1) what is the evidence for psychological trauma following surgery in children and their parents, and 2) what factors are associated with trauma responses in the same groups.

## Methods

### Search strategy and selection criteria

We conducted a systematic review with a separate meta-analysis with an overall summary estimate from available data. We searched the literature using PubMed, Web of Science and PsycInfo databases.

We used the following search terms: *(Psychological distress OR anxiety OR depression OR mental OR trauma OR traumatic OR post-traumatic stress OR acute stress) AND (children OR adolescent OR family OR sibling OR parent OR carer OR paediatric OR pediatric) AND (surgery OR surgical OR operation OR hospital OR procedure OR anaesthetic)*. Searches were performed in February 2021 by two independent reviewers who then conferred to agree inclusion and exclusion.

We also performed a separate search using Google Scholar and by searching references of included texts. Google Scholar searches were conducted using the search criteria “PTSD surgery children” and “Acute stress disorder surgery children”. We also screened reference lists of included studies for additional papers missed in initial searches.

A priori inclusion criteria were: 1) studies which reported on psychological trauma in children undergoing surgery; 2) studies which reported on psychological trauma in parents of children undergoing surgery; 3) observational or intervention studies. Exclusion criteria were 1) studies which did not use a validated measure; 2) studies which reported other mental health variables if there was not an explicit measure of trauma; 3) literature reviews other than systematic reviews.

### Data analysis

Studies were extracted to Endnote which was used to remove duplicates. Quality of studies was assessed using the Newcastle-Ottawa Quality Assessment Scale, adapted for use with cross-sectional studies.^19^ Studies can achieve an overall rating of a maximum of 10 stars. Each study in the review was assessed by one researcher (SK) scored appropriately. A selection of studies were blindly assessed by another researcher (DT), with ratings compared and found to match.

### Meta-analysis

Prevalence rates reported in studies for PTSD were pooled in meta-analysis using STATA (version 16) for studies reporting prevalence in 1) children and 2) parents. The metaprop command was utilised, with a random effects model using the Freeman-Tukey double arcsine transformation was used, with pooled estimates and 95% confidence intervals reported.^20^ Sensitivity analyses were performed by removing studies with greatest bias (<5 total NOS score). Where available, data was extracted from individual studies to perform meta-regression to identify potential moderator variables on affect sizes across pooled studies.

## Results

Initial searches yielded a total of 1,671 papers, 609 of which were identified as being duplicates and subsequently removed. The abstracts of the remaining 1,062 papers were examined for relevance. In total, 149 papers were then retrieved and reviewed for inclusion. A total of 16 papers were included in the final review. Figure A shows a summary of the search and study selection process. Table One provides a summary of the studies included.

**Figure A.**
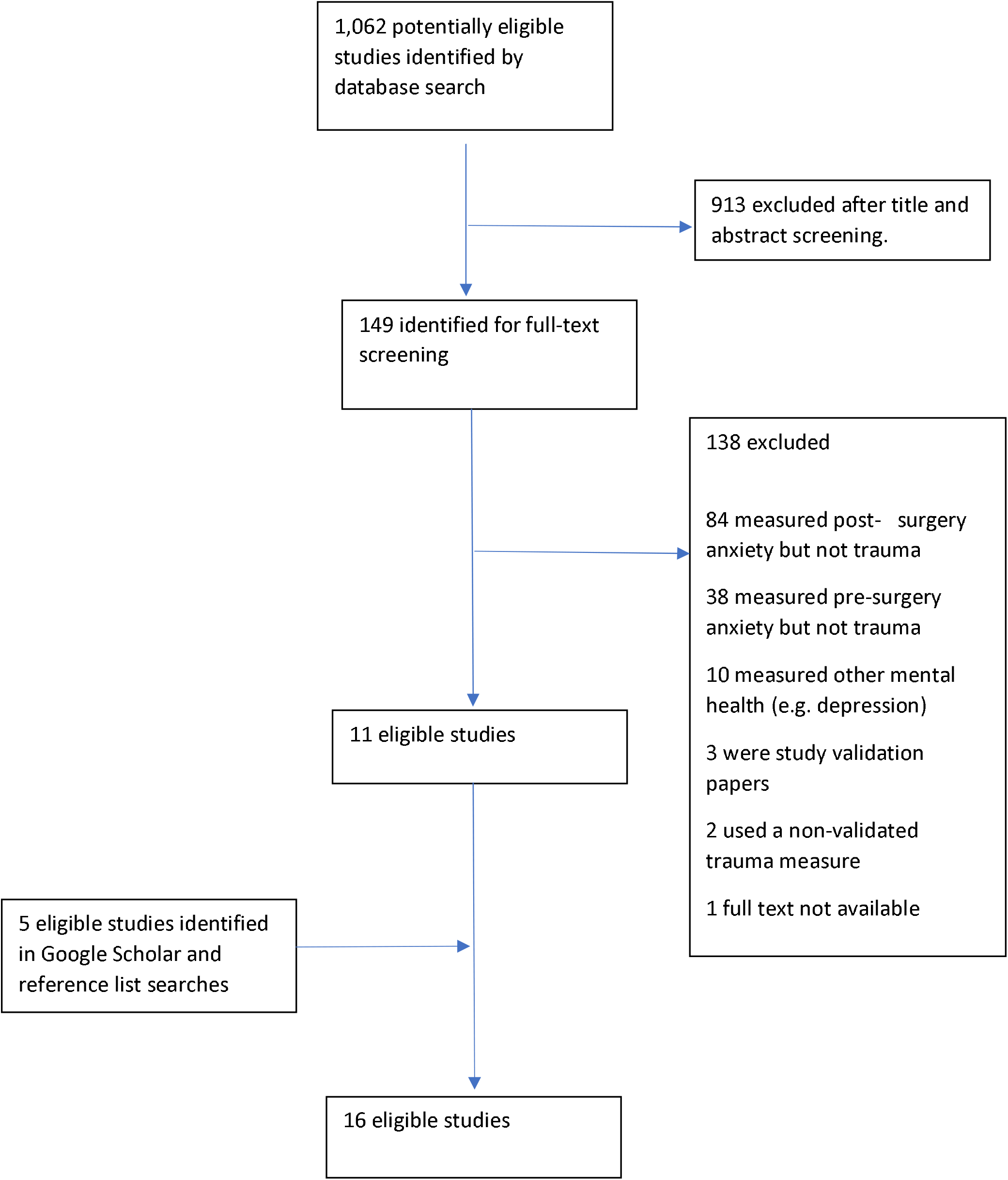
Study selection flowchart

**Table one.**
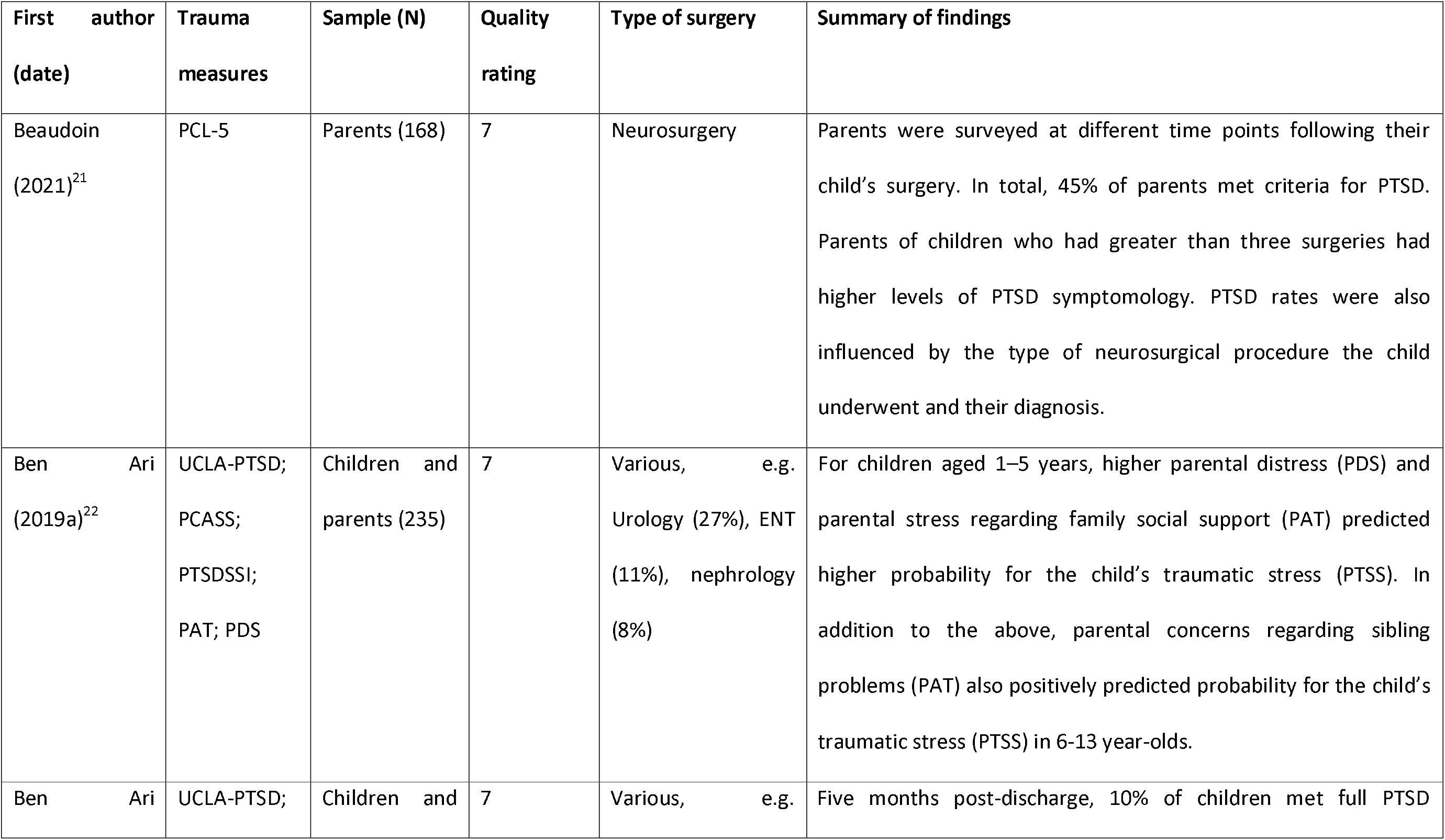
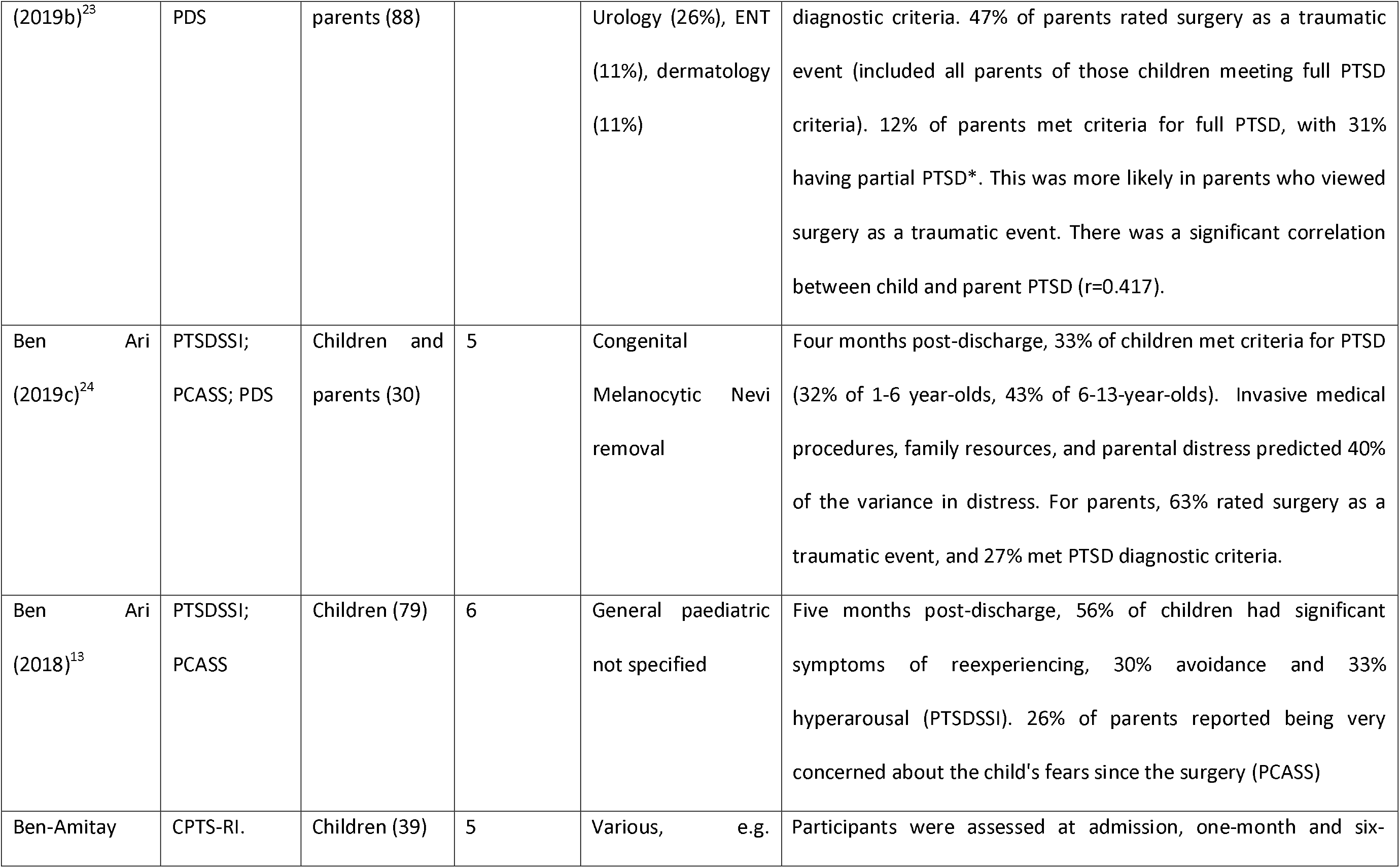
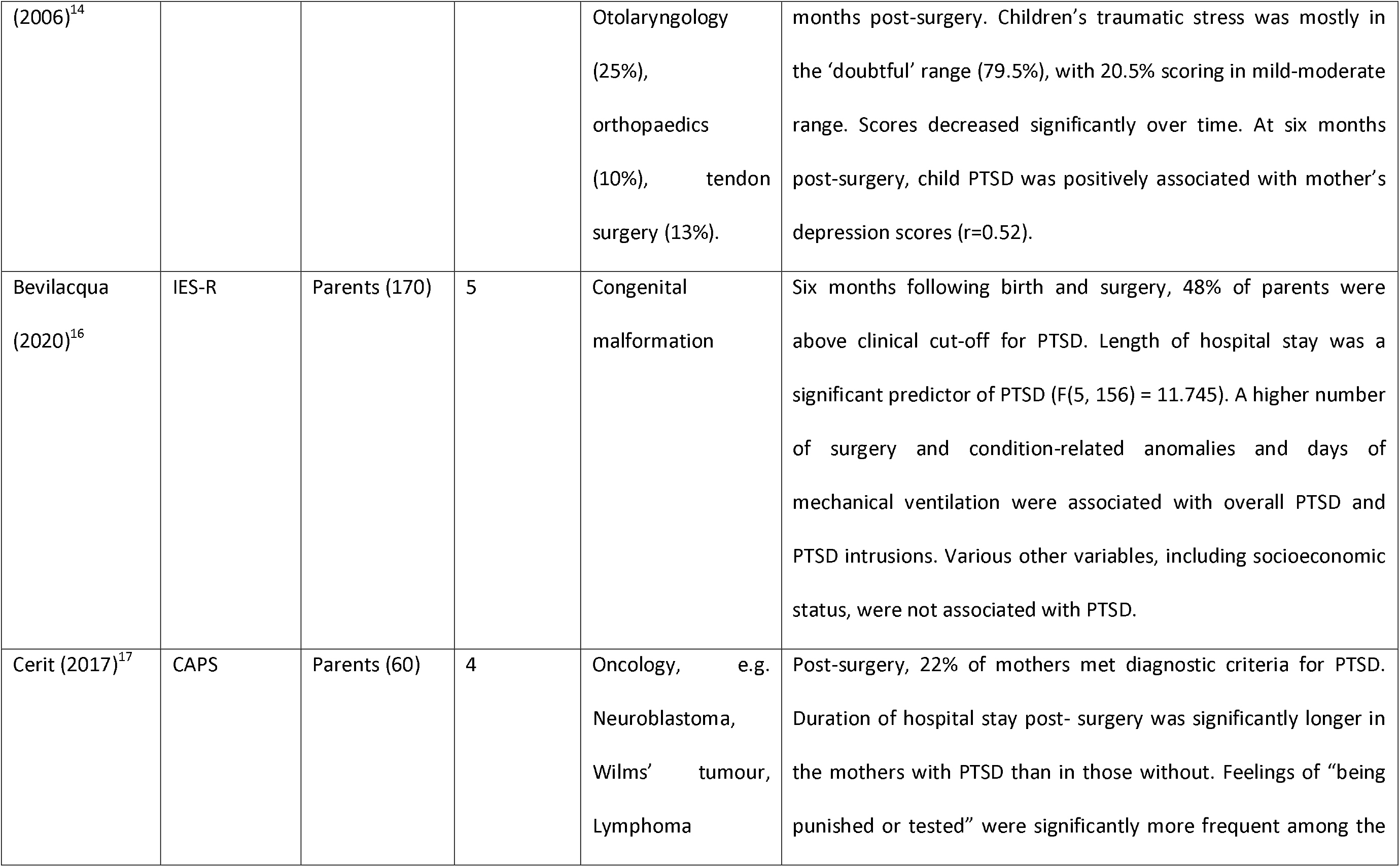
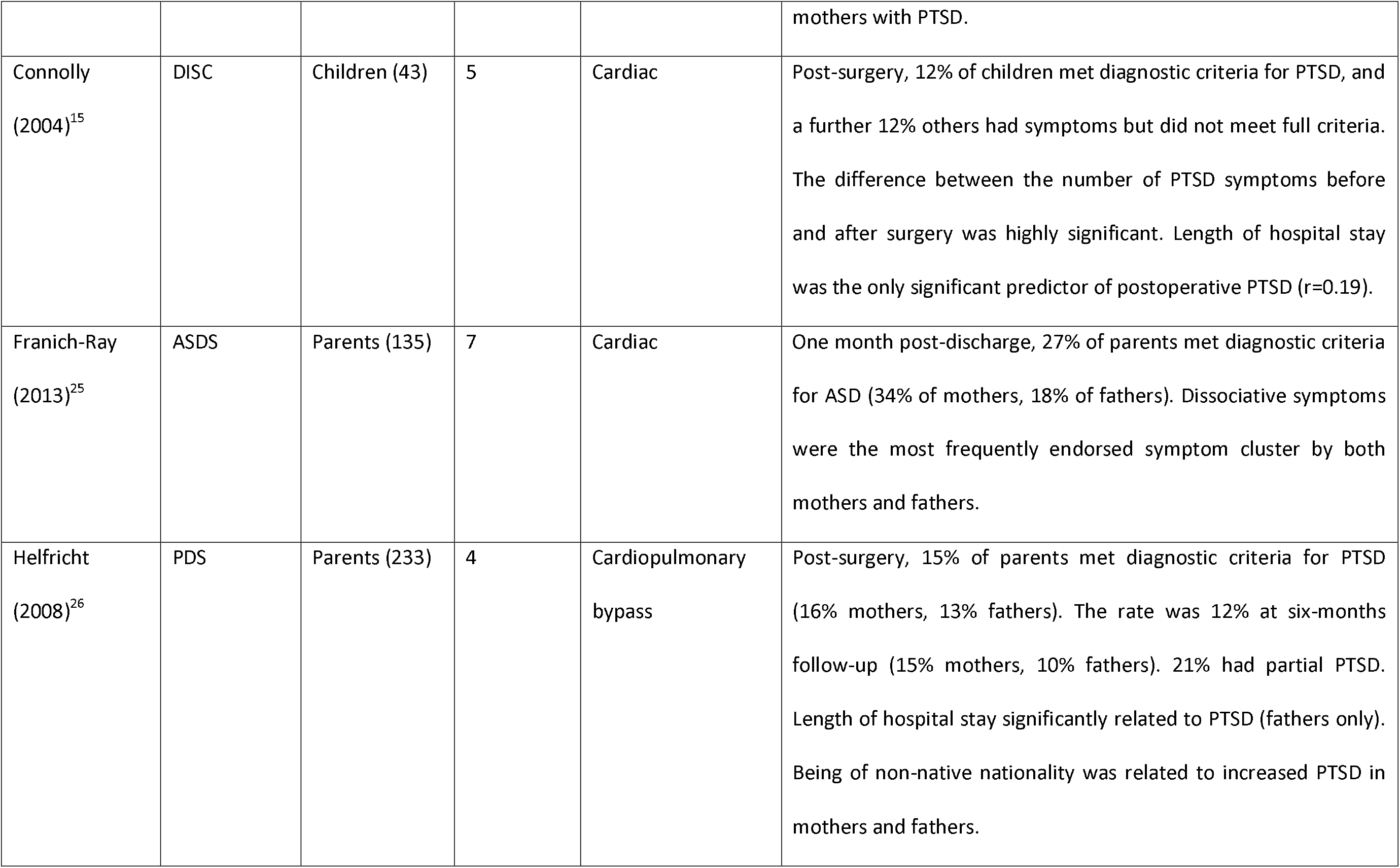
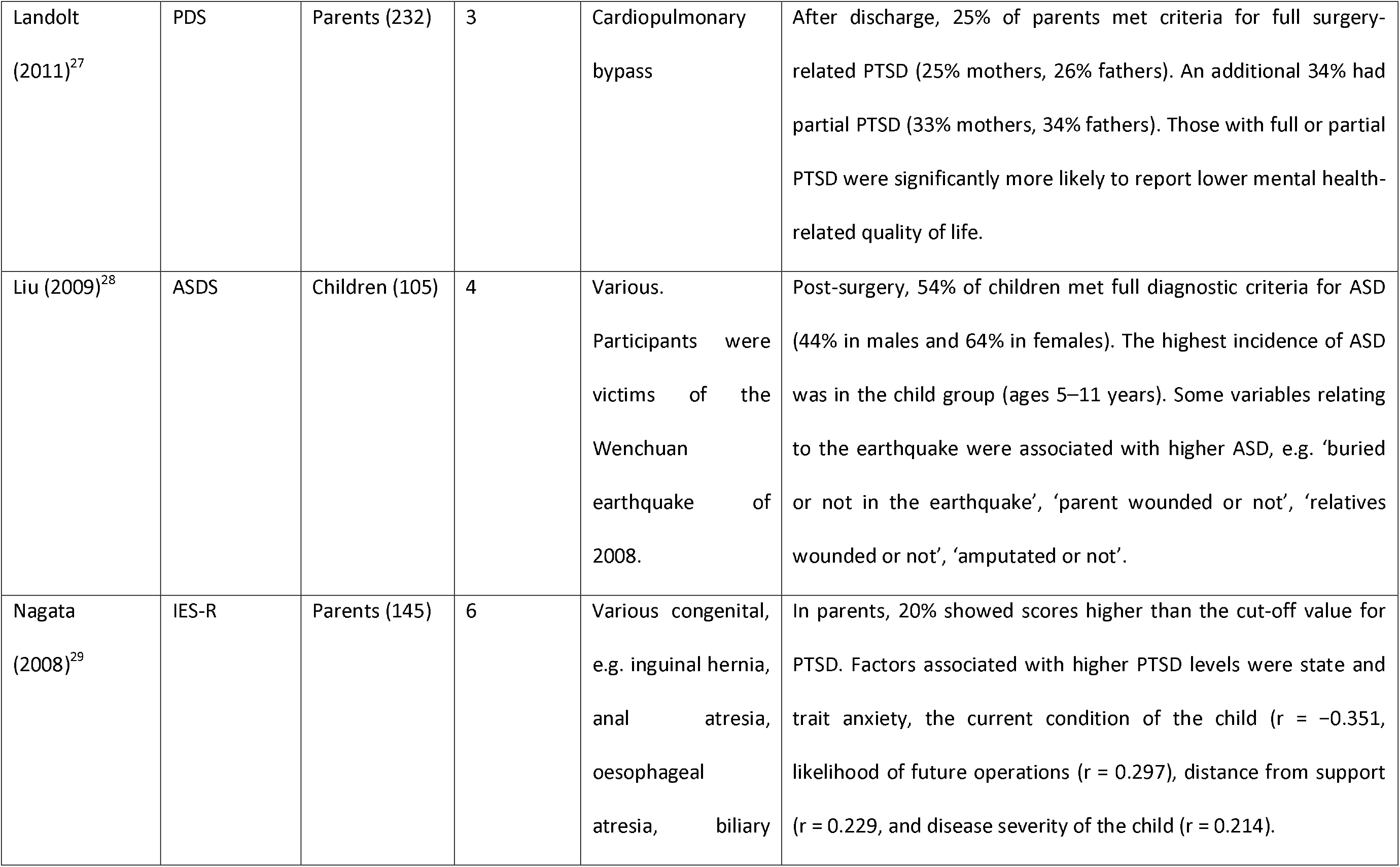
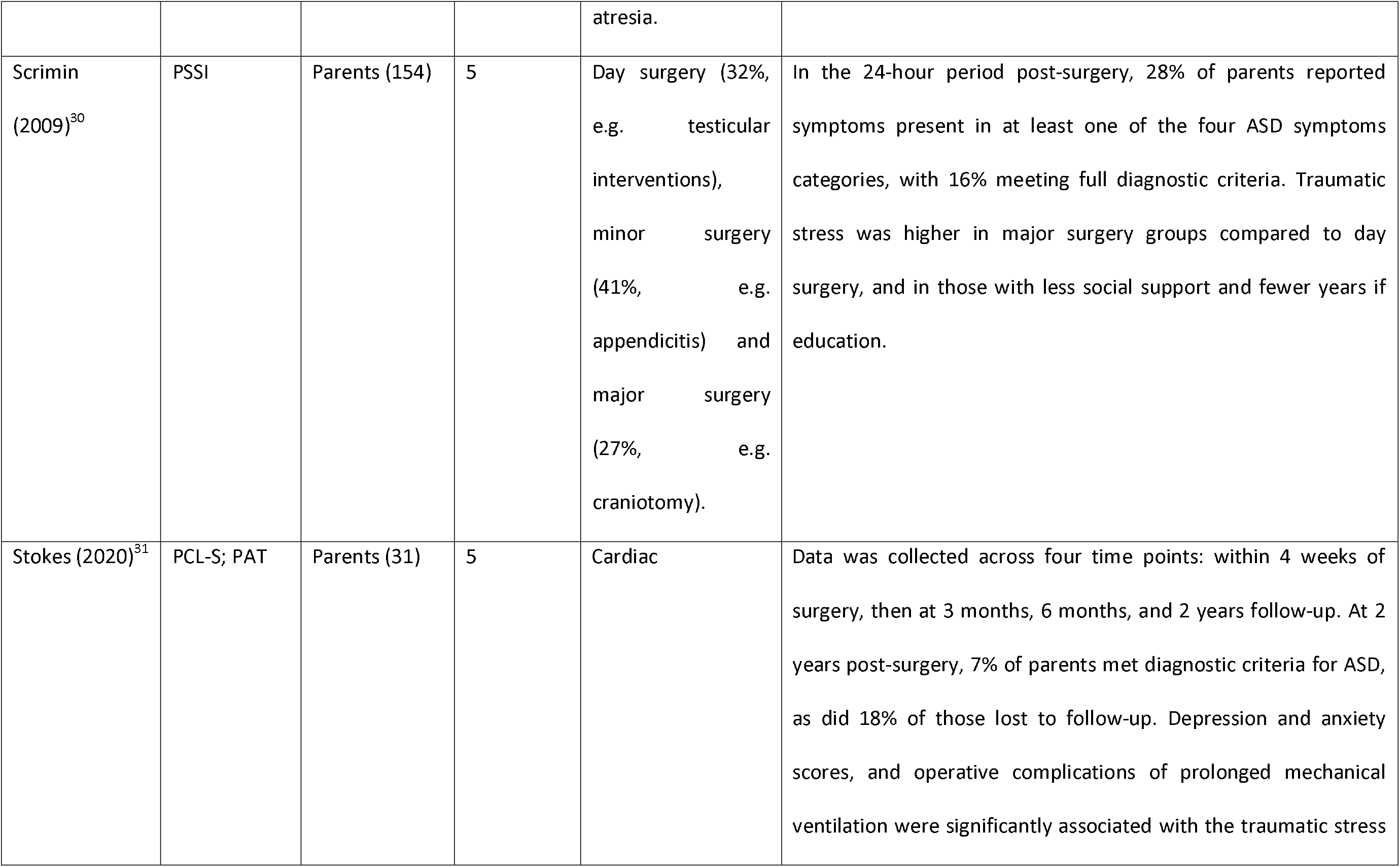
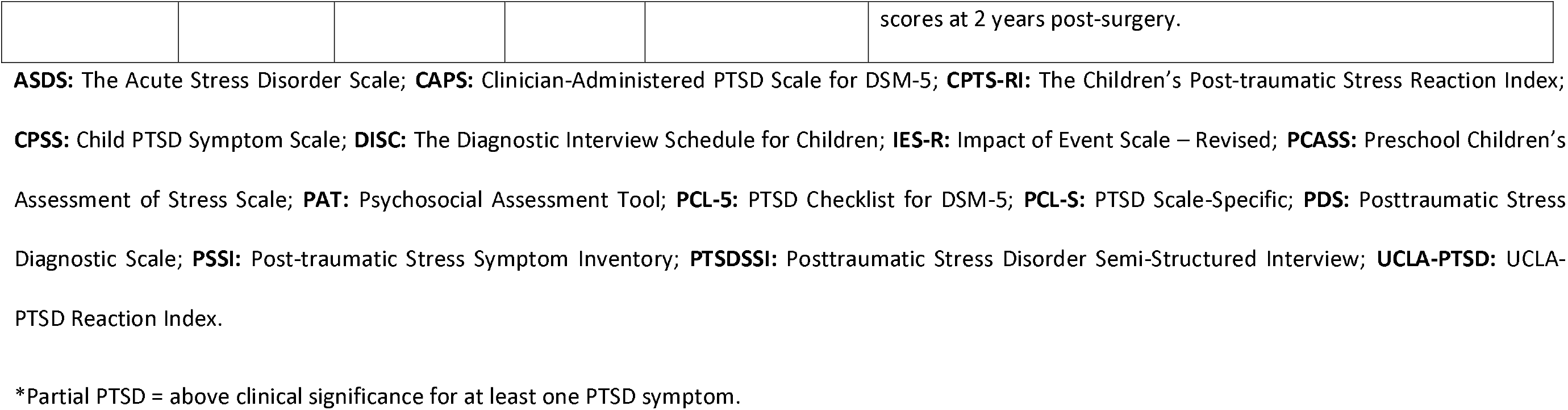
Overview of studies.

There was variation in the types of surgery involved in each study, with the most common being cardiac (5 studies). In total, six (38%) studies investigated traumatic stress just in child participants, while eight (50%) studies reported data on parents only. A further two (12%) studies reported on both children and parents concurrently.

Most studies (13) reported whether participants met diagnostic or clinical thresholds for PTSD or ASD, as per the cut-off ranges of the respective measures used. One study gave a breakdown of PTSD symptoms without reporting the number of participants who reached threshold for diagnosable PTSD. The majority of studies (12) provided data regarding correlating or risk factors associated with higher symptoms of traumatic stress.

### Prevalence findings in children and young people

In total, six studies suggested that at least some children had developed PTSD or ASD symptoms after surgery.^13-15,23,24,28^ One study by Liu and colleagues^28^ included participants who had been involved in a natural disaster prior to surgery, which was not controlled for, and may have inflated rates of traumatic stress. Another study did not report overall rates of traumatic stress, but did report that high numbers (56%) of children displayed difficulties with specific PTSD symptoms, e.g. flashbacks^13^.

The four remaining studies which reported overall prevalence of PTSD symptoms at least 3 months post-surgery were pooled in meta-analysis (see Figure B) for a total of 187 children. All included studies had a NOS score >= 5. Heterogeneity was high (I^2^ = 80%). Overall pooled estimate of prevalence was 16% (95% CI 5-31%). Available data from studies allowed meta-regression of type of surgery (cardiac versus other), mean age of children, and sex proportion in samples as predictors of prevalence, with none showing significant association (data not shown).

**Figure B :**
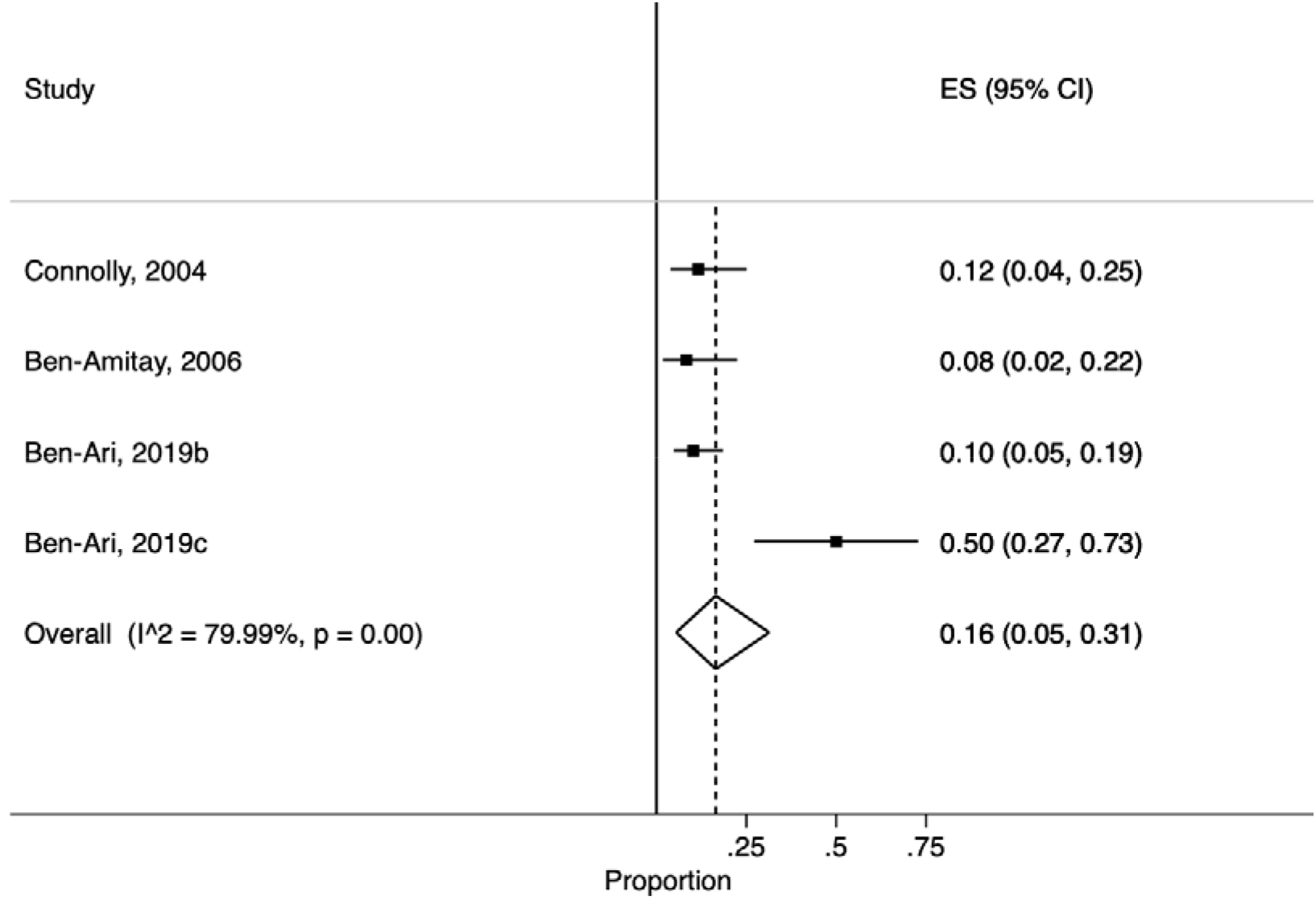
Forest plot of proportions of children meeting criteria for PTSD > 3 months after surgery with pooled estimates of overall prevalence (with 95% confidence intervals).

### Prevalence findings in parents

In total, 11 studies reported rates of traumatic stress symptoms in parents.^16,17,21,23-27,29-31^ All of these studies reported that at least some parents self-reported symptoms consistent with full PTSD diagnostic criteria. The 11 included studies which reported prevalence of PTSD symptoms at least 3 months post-surgery in parents were pooled in meta-analysis for a total of 1444 parents from all studies (see Figure C). Heterogeneity was high between studies (I^2^ = 91%). Overall pooled estimate of prevalence of PTSD was 23% (95% CI 16-31%). Removal of studies with lowest NOS score did not improve heterogeneity, or affect overall pooled estimate significantly. Available data from studies allowed meta-regression of type of surgery, mean age of children and parents, sex proportion of children and parents, and parental further education level as predictors of prevalence, and none were

**Figure C:**
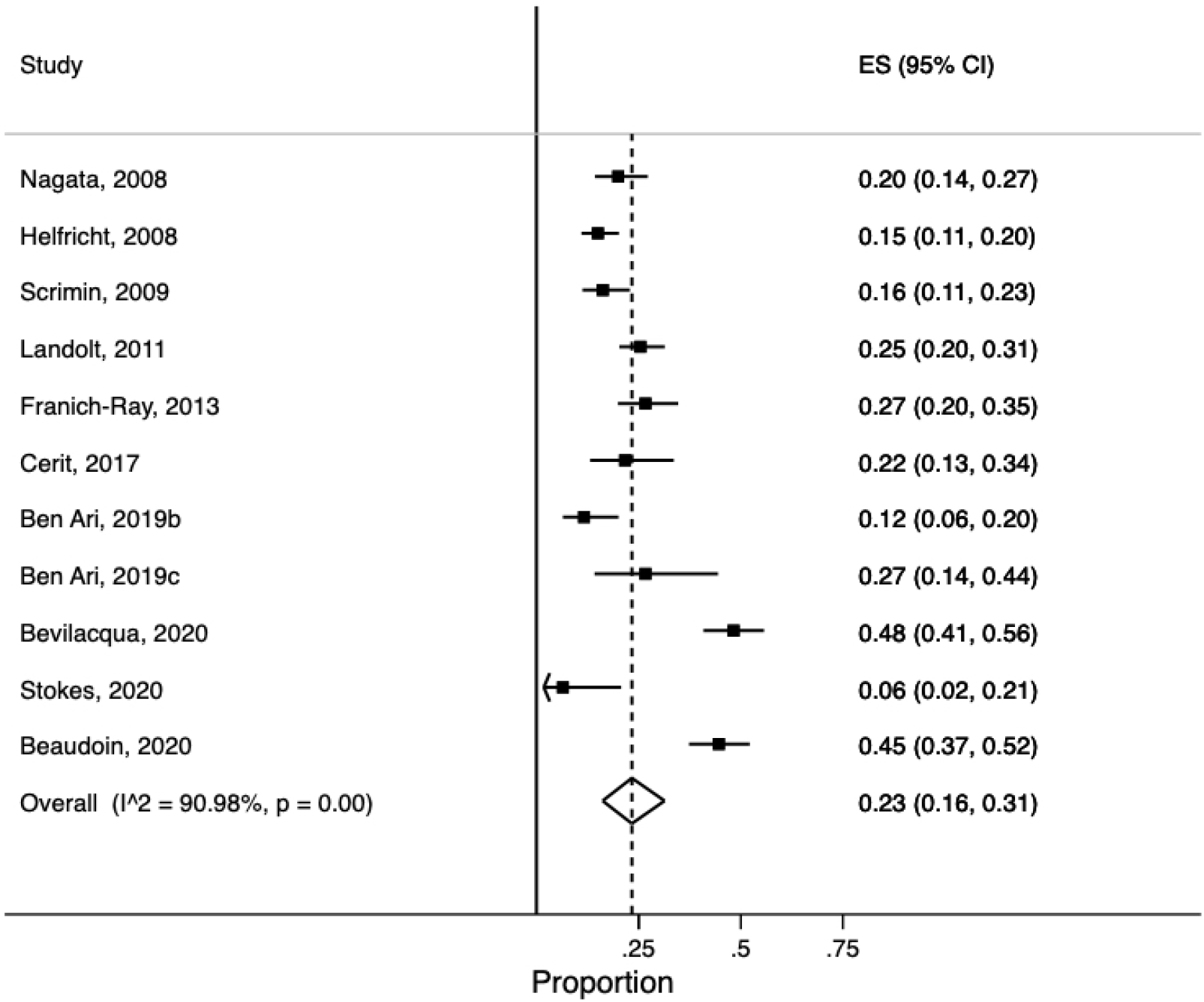
Forest plot of proportions of parents meeting criteria for PTSD > 3 months after surgery with pooled estimates of overall prevalence (with 95% confidence intervals)

### Findings on potential influencing factors within studies

Several studies conducted additional analyses to investigate the presence of factors which were associated with or predicted PTSD development in children and parents. A number of variables were reported. Four studies showed that length of stay in hospital was positively associated with PTSD,^15-17,26^ Some authors^17^ suggested that this finding was explained by length of stay in hospital reflecting illness severity in the child. Others proposed that children who spend longer in hospital will be exposed to higher levels of accumulative trauma.^15^ This assumes the entire hospital experience to be traumatic, including associated procedures and recovery, over and above the surgery itself. One study which just found this association in fathers, suggested that the longer the hospital stay, the more parents may struggle to meet the demands of work and family commitments whilst the child remains in hospital.^26^ This explanation would suggest that the depletion of resources contributes to traumatic stress development, or that parents who cannot work due to the caring for a hospitalised child may face financial or other burdens which increase stress and decrease ability to cope.

It is likely that a combination of factors relating to the duration of hospital stay during and post-surgery contribute to the development of traumatic stress in children and parents. A consistent finding, this risk factor is easily recorded and could be used to identify those more in need of additional psychological support.

Several studies reported that different factors relating to the severity of the surgery, complications within the surgery, or severity of the child’s illness can contribute to the development of traumatic stress. For example, studies reported that more invasive procedures,^24,30^ complications such as the requirement for mechanical ventilation,^16,31^ or the severity of the child’s condition^29^ were all associated with higher PTSD. These findings related to both children and parents.

In three (19%) studies, associations were found between parental mental health and child traumatic stress.^14,23,24^ Specific difficulties that parents reported were PTSD and depression. These findings suggest that the distress and ability to cope of parents can play a significant role in the mental health and traumatic stress of children following surgery.

The direction of these associations is not necessarily known and could feasibly operate in either direction, i.e. parental mental health leads to children being more likely to develop traumatic stress, or where children develop traumatic stress, parents are more likely to suffer with their own mental health. These findings are potentially important in helping to identify those who may benefit from additional support.

Some factors relating to social and family support appear to be important when considering traumatic stress following surgery. One study found that when lower levels of family social support were reported using the Psychosocial Assessment Tool II,^32^ children were more likely to report traumatic stress.^22^ In this study, social support included psychological help, financial support, daily help, and help with childcare. Authors suggested that a lack of such resources made it more difficult for children to cope with the stress of the surgery and hospitalisation, not least because it might mean parents were less available to support them. In another study, parents had higher levels of traumatic stress if they reported being far away from their support network, or had fewer social contacts around the time of the surgery.^29,30^ Additionally, one study found that being a non-native nationality was related to symptom severity and that mothers who used the social services provided by the hospital presented with more severe symptoms of acute PTSD.^26^ Collectively, these findings suggest that social support is an important factor in the development of surgery-related traumatic stress in both children and parents.

## Discussion

We believe that this is the first systematic review and meta-analysis of the literature examining post-traumatic stress reactions in children and parents, following surgical intervention in a child. In meta-analysis, pooled estimates of prevalence of post-traumatic stress was 16% in children, and 23% in parents. Past epidemiological research has suggested that 8% of children in the general population will develop PTSD by 18 years, a figure which rises to 25% in those who experienced at least one trauma in their lives.^33^ Lower prevalence figures have been reported for adults who have experienced trauma.^34^ This review therefore suggests that paediatric surgery may lead to an increase the risk of PTSD, which has implications for screening and service provision for children undergoing surgery, and their families.

We found evidence that a range of psychological and social factors might increase the risk of post-traumatic stress within studies. We did not find evidence for moderating variables in meta-regression once studies were pooled, though there was inconsistent reporting of potential influencing variables across studies thus potentially limiting power. The most commonly reported risk factor in studies was the length of time spent in hospital. This may be explained either by those being in hospital for longer being more exposed to potentially traumatic events, are more unwell, or have had a more serious surgical intervention. It is likely that patients who have experienced complications or multiple procedures are also in this group.

Research suggests that individuals with more social support are less likely to develop chronic PTSD symptoms.^35-40^ There was some evidence to support this in the current review, with several studies reporting a lack of family and social support being associated with traumatic stress in both children and parents. These findings may also relate to duration of hospital stay, in that children who spend longer in hospital are more likely to miss out on support from extended family or friends, or parents who spend significant amounts of time visiting or staying with a child in hospital may be less able to access social support during this period.

Our search had a number of strengths. The literature search was systematic and robust, including rigorous searches of multiple databases and sources. A priori search terms were used, with agreed inclusion and exclusion criteria for selecting suitable studies. Throughout the search, two researchers were used to cross-reference studies in terms of the inclusion/exclusion criteria, as well as is the quality rating of studies. This will have reduced the risk of error and increased reliability. Our review was necessarily narrow in focus; investigating post-traumatic stress reactions solely. However, children and parents may well present with other mental health (e.g. anxiety) or behavioural concerns following surgery; the investigation of which was beyond the scope of this review. In young children particularly, indicators of post-traumatic stress may manifest in ways that are not captured in standard trauma measures, such as via behavioural difficulties. It is important therefore, that services consider wider aspects of mental health, behaviour and well-being when considering the possible impact of paediatric surgery.

The interpretation of findings is limited by the cross-sectional design of studies, meaning that firm conclusions about the cause of traumatic stress in patients are hard to draw. For example, we found that duration of hospital stay was associated with higher levels of traumatic stress. However, there are several associated variables that might explain this, such as whether patients perceive their situation to be life-threatening,^24,30,41-43^ which might be more likely in children who spend longer in hospital as they are more likely to have undergone more serious surgery, or experienced complications. Further, it was not possible to separate the impact of the surgery specifically, from the incident or illness which led to the child requiring surgery, which may in if itself be experienced as traumatic, e.g. an accident leading to injury. None of the studies we found controlled for this issue.

None of the studies in the review included a measure of traumatic stress at baseline (before surgery) and again post-surgery. This is important as a potential source of bias, since the absence of a baseline measure makes it hard to conclude whether the surgery directly preceded the onset of traumatic stress reactions, or whether traumatic stress was already present prior to surgery. Additionally, no studies in the review included control groups against which levels of traumatic stress could be compared. Because we found a number of studies reporting proportions, we were able to perform a meta-analysis. The heterogeneity was high, however we did use a random effects model and performed sensitivity analyses by removing studies with the greatest bias.

Many studies did not have long-term follow-up and only measured traumatic stress directly following surgery. Some research suggests that as many as 56% of individuals who develop PTSD will spontaneously recover within five months of onset^10^ and so findings in the short term may not have been maintained. None of the studies included details of whether participants received appropriate interventions for traumatic stress which may have influenced measures at follow-up. Future research in this area should include follow-up measurements to improve our understanding of the long-term persistence and effects of traumatic stress post-surgery.

Future studies should include baseline, pre-surgery measures of trauma symptoms, plus traumatic event checklists where participants are asked to report types of traumatic events they have experienced. This will help to clarify the role of surgery in the development of traumatic stress reactions. Our study provides evidence of sufficient levels of traumatic stress following paediatric surgery to warrant the evaluation of interventions to support children and families where the child undergoes surgery. Clinical guidelines recommend a period of *active monitoring* for mild symptoms of a period of four weeks, as a way of supporting individuals in the weeks following a trauma.^4^ Future research could investigate report on the implementation of such interventions and on their effectiveness.

## Conclusions

There is consistent evidence in the literature of post-traumatic stress following surgery in childhood which warrants further investigation. Cases where the surgery is more complex, or the associated health condition is serious or life-threatening, appear to increase the risk of traumatic stress reactions. Several other risk factors have been identified, which may guide services in screening those who may be most at risk. These include cases where the child is in hospital for a longer period, the family are lacking in social support, or a parent is suffering with mental health difficulties or high levels of stress.

Those delivering surgical care to children would benefit from a raised awareness of the potential for post-traumatic stress in their patients and their families, and offer screening and support. This is particularly relevant in case of service disruptions like during the current pandemic, where screening for traumatic stress and other mental health difficulties is challenging. Those interventions may provide support to those who report difficulties or are more at risk.

## Data Availability

This is a meta-analysis but data extracted from studies used is available

